# Hierarchical Two-Stage Cost-Sensitive Clinical Decision Support System for Screening Prodromal Alzheimer’s Disease and Related Dementias

**DOI:** 10.1101/2022.09.06.22279650

**Authors:** Michael J. Kleiman, Taylor Ariko, James E. Galvin, the Alzheimer’s Disease Neuroimaging Initiative

## Abstract

**Background:** The detection of subtle cognitive impairment in a clinical setting is difficult, and because time is a key factor in small clinics and research sites, the brief cognitive assessments that are relied upon often misclassify patients with very mild impairment as normal. In this study, we seek to identify a parsimonious screening tool in one stage, followed by additional assessments in an optional second stage if additional specificity is desired, tested using a machine learning algorithm capable of being integrated into a clinical decision support system.

**Methods:** The best primary stage incorporated measures of short-term memory, executive and visuospatial functioning, and self-reported memory and daily living questions, with a total time of 5 minutes. The best secondary stage incorporated a measure of neurobiology as well as additional cognitive assessment and brief informant report questionnaires, totaling 30 minutes including delayed recall. Combined performance was evaluated using 25 sets of models, trained on 1181 ADNI participants and tested on 127 patients from a memory clinic.

**Results:** The 5-minute primary stage was highly sensitive (96.5%) but lacked specificity (34.1%), with an AUC of 87.5% and DOR of 14.3. The optional secondary stage increased specificity to 58.6%, resulting in an overall AUC of 89.7% using the best model combination of logistic regression for stage 1 and gradient-boosted machine for stage 2.

**Conclusions:** The primary stage is brief and effective at screening, with the optional two-stage technique further increasing specificity. The hierarchical two-stage technique exhibited similar accuracy but with reduced costs compared to the more common single-stage paradigm.

## INTRODUCTION

Alzheimer’s disease and related dementias (ADRD) may be difficult to detect in clinical research and general clinical practice, especially in prodromal stages (i.e., mild cognitive impairment or MCI) where cognitive changes are subtle and can be mistaken for normal aging [1]. Examining biomarkers characteristic of ADRD such as amyloid or phosphorylated tau via spinal fluid, blood tests, or PET scans support the presence of underlying pathology without specifically determining current cognitive status of the individual [2]. Routine screening for MCI and ADRD to detect early impairment is not commonly utilized in primary care due to a number of factors including time and effort, challenges with administering and interpreting brief cognitive tests, and lack of screening guidelines [3–5]. Similar challenges exist for screening in clinical trials, especially for MCI cases.

While a recent approval of therapeutics that seek to delay onset or slow progression has been controversial [6], symptomatic medications can delay disease progression and mortality [7] and nonpharmaceutical and lifestyle interventions may provide cognitive or functional benefits [8,9]. Other benefits of screening include the ability to change behaviors or improve health outcomes [10] and advanced care planning. Thus, screening for and early detection of MCI and ADRD has the potential to offer clinical benefit today, and the development of effective programs may enhance clinical research and patient selection for emerging disease-modifying medications in the future.

Brief cognitive assessments, including the Montreal Cognitive Assessment (MoCA) [4] and Mini-Mental State Exam (MMSE) [11] are effective at identifying cognitive impairment, however they may not be as sensitive for MCI, particularly non-AD forms [5]. Self-report screening instruments, including the Quick Dementia Rating Scale (QDRS) [12], Everyday Cognition Scale (ECog) [13], and Functional Activities Questionnaire (FAQ) [14], can identify subjective complaints and early changes in instrumental activities of daily living (IADL). Given sociodemographic and educational biases inherent in many cognitive assessments, global screening measures such as the QDRS and ECog may be more sensitive to earlier stages of impairment [15] and provide measures of changes in cognitive and functional abilities over time [16]. However, the self-report and informant-report measures are subjective, and cannot be used on their own to determine any objective measure of cognitive performance [17]. As a result, screening for cognitive impairment may benefit from incorporating both an objective cognitive assessment component as well as self-report and/or informant-report measures, to better identify individuals with early impairments and reassure individuals with a low likelihood of cognitive impairment.

Because clinical practices and research centers outside large tertiary academic medical centers may not have the available time, effort, and or trained staff to conduct comprehensive cognitive evaluations, there often is an overreliance on these brief screening measures that may potentially miss detection of up to half of true cases of cognitive impairment [18,19]. This can have consequences on clinical care and on referral for clinical trials. To address these unmet needs, two strategies can be utilized: automating interpretation, and simplifying the assessments.

### Automating interpretation using clinical decision support systems

Clinical decision support (CDS) systems help health care professionals by performing various functions including giving reminders, interpreting tests, assisting diagnosis, and alerting of medication interactions. CDS systems can effectively reduce healthcare costs, for example by reducing unnecessary laboratory testing [20] or aiding physicians in differential diagnosis [21]. Studies implementing CDS systems have demonstrated improved diagnostic accuracy and documentation, as well as reduced diagnostic error [21,22]. CDS systems can also improve screening for common chronic diseases such as cancer, kidney disease, obesity, abdominal aortic aneurysm, diabetes, osteoporosis, hepatitis B virus, depression, and dementia, leading to improved rates of diagnosis [23,24]. There is clear potential for CDS systems to help close the gap between healthcare provider knowledge and performance for a more robust clinical decision-making system.

Early and accurate detection of ADRD is vital to early case ascertainment and recruitment for clinical trials and can be aided by CDS systems. Studies of CDS systems’ effectiveness at detecting dementia in primary care identify significant improvements in rates of reported dementia cases [25] as well as physician confidence in differential diagnosis [26], compared to when the CDS was not utilized. Machine learning is also useful in CDS systems for feature selection as well as model development by optimizing model inputs and allowing for complex data relationships in modeling [27]. A review of the contribution of machine learning in classification of MCI and ADRD using the Alzheimer’s Disease Neuroimaging dataset reported overall improvement in classification and prediction accuracy, especially in challenges involving MCI patients [28]. Furthermore, a study using a machine learning-based dynamic CDS system for supporting the diagnosis of dementia achieved an excellent classification accuracy of 92% [29].

### Cost-sensitive cognitive screening

In addition to increasing accuracy, the main benefit of CDS systems is to decrease monetary and time costs associated with screening and subsequent diagnosis. Many CDS systems scour electronic medical records (EMR) for treatment regimens, physician notes, and/or the patient’s medical history. These datapoints are able to identify patients who may be at risk for a particular disorder with sufficient accuracy [30,31], however these systems require the EMR to contain sufficient detail to determine that risk. In the case of MCI and ADRD, studies that use EMR for risk assessment often rely heavily on comorbidities that indicate poor health, which then secondarily indicates risk of ADRD. No method for identifying latent factors of cognitive impairment itself within EMR, and not just determining risk factors, has been successful to date. Thus, to properly screen for prodromal impairment, components that directly assess cognitive and daily functioning must be incorporated into the medical record [32].

Many brief cognitive screeners, including the MoCA [4] and MMSE [11], require licensing and training for use, and can misclassify individuals with very mild impairment as not impaired [5], particularly individuals from underrepresented and underserved communities. While combining these brief cognitive screeners with a self-report screener such as the QDRS or FAQ can improve overall detection accuracy, producing a classification of “screen positive” or “screen negative” based on the results of multiple tools introduces a degree of subjectivity on the part of the clinician doing the interpretation.

Cost-sensitive CDS systems have also been shown to greatly improve screening accuracy while minimizing assessment time [33,34]. To minimize time cost, the components examined are carefully curated using feature selection machine learning algorithms [35], which serve to identify the most useful features within a given set of assessments. In previous work, four assessment components were found to detect impairment at 94.5% sensitivity within 15 minutes of active clinician time: delayed narrative recall, trail-making B, and memory questions reported by both the patient and an informant [36]. These components were also found to be similarly highly valued in other feature selection studies, highlighting the utility of patient and informant reported measures [33] as well as the benefit of both a delayed memory component [34,37] and executive-visuospatial component [37]. Other studies have placed greater focus on minimizing costs over measuring cognitive performance, determining that self-report questions alone can produce an impressive area under the ROC curve (AUC) of 0.865 when classifying a patient using the Clinical Dementia Rating (CDR) scale [33]. However, applying these particular neuropsychological tests in routine clinical practice may have practical and cost limitations.

### Multi-stage screening

In this study, we evaluated the combined efficiency of cost-sensitive screening and automated interpretation with the efficacy of robust assessments by developing a multi-stage screening paradigm capable of being integrated into CDS systems for use in primary care and research. In contrast to a single stage, two or more hierarchical stages enable easily-collected screening assessments (e.g., questionnaires, brief assessments) to be first examined prior to those that require more time and effort to collect (e.g., neuropsychological testing, MRI). If sensitivity is prioritized to minimize false-negatives, patients that screen negative at early stages could be safely excluded, and only those that are not clearly unimpaired would be recommended for additional screening procedure. This technique could empower smaller clinics and research sites to screen for cognitive impairment without expending unnecessary resources, and without requiring physicians to interpret disparate screening procedures. Patients who screen positive could then be further evaluated or referred to memory-care specialists for further diagnosis and management.

We hypothesize that a two-stage screening paradigm, that uses progressively more in-depth screening components at each stage, will exclude more non-impaired patients after the final stage and misclassify fewer impaired patients overall than if only a single-stage algorithm was used on all patients. We aim to develop a parsimonious and brief primary stage that effectively screens early impairment, with an optional secondary stage that further excludes healthy participants while using more time-intensive and costly assessments. More in-depth diagnostic evaluation could then be recommended to more accurately identify impairment status, determine dementia etiology, and prompt management and/or treatment of MCI and ADRD when required.

## METHOD

### Participants

Two sets of participant data were used in this study: a research dataset for training and parameter optimization, and a clinical dataset for testing and providing output statistics.

Obtained from the *Alzheimer’s Disease Neuroimaging Initiative* (ADNI), 1181 participants (517 HC, 522 MCI, 142 AD) were used to train each set of models as well as in feature selection and hyperparameter optimization. The ADNI database (adni.loni.usc.edu) was launched in 2003 as a public-private partnership, led by Principal Investigator Michael W. Weiner, MD. The primary goal of ADNI has been to test whether MRI, PET, other biological markers, and clinical and neuropsychological assessment can be combined to measure the progression of MCI and early AD. While subjects in ADNI are voluntary research participants and thus are less diverse and more highly educated than those found in the general population, this data was useful as a training set as its participants are examined at dozens of testing centers across the globe yet are each administered roughly identical assessment regimens.

To test the models, 127 participants (41 HC, 59 MCI, 27 AD) from the *Comprehensive Center for Brain Health* (CCBH) were used. Unlike ADNI, the CCBH dataset is drawn from a clinical population with a focus on examining brain health, MCI, and ADRD. As a result, the makeup of the CCBH cohort is more representative of populations of real-world clinics and medical practices than the ADNI dataset despite having similarly limited racial and ethnic diversity, and thus serves as an effective group to test models for clinical decision support. The two datasets share many common features, with the main exception being that patient self-report of cognition and functioning is captured by the ECog in ADNI and by the QDRS in CCBH.

From both datasets, only participants with a CDR of 0 (no dementia), 0.5 (questionable or very mild dementia), or 1 (mild dementia) were selected, ensuring that the participants resemble those who would seek screening procedures for cognitive impairment. Those with moderate to severe dementia (CDR 2 or 3) can be more readily identified as impaired, and thus were excluded from our screening procedure.

### Two-stage architecture

The CDS system developed in this study is intended to primarily select out healthy controls, and identify ideal candidates for cognitive impairment screening, while minimizing time and effort costs. To achieve this, the system is given a two-stage hierarchical structure, where the primary stage is intended to screen out those with no impairment and the second stage is intended to further improve specificity after introducing additional assessments. While the primary stage thus prioritizes sensitivity over specificity, the second stage utilizes a multi-model network [38], which allows for separate models each with their own set of features and hyperparameters to target either the impaired or nonimpaired class. This strategy enables the ability to fine-tune each model’s performance, as well as set individualized classification thresholds, to optimize the detection of impairment status. A visualization of the architecture can be seen in **Figure 1**.

**Figure 1.**
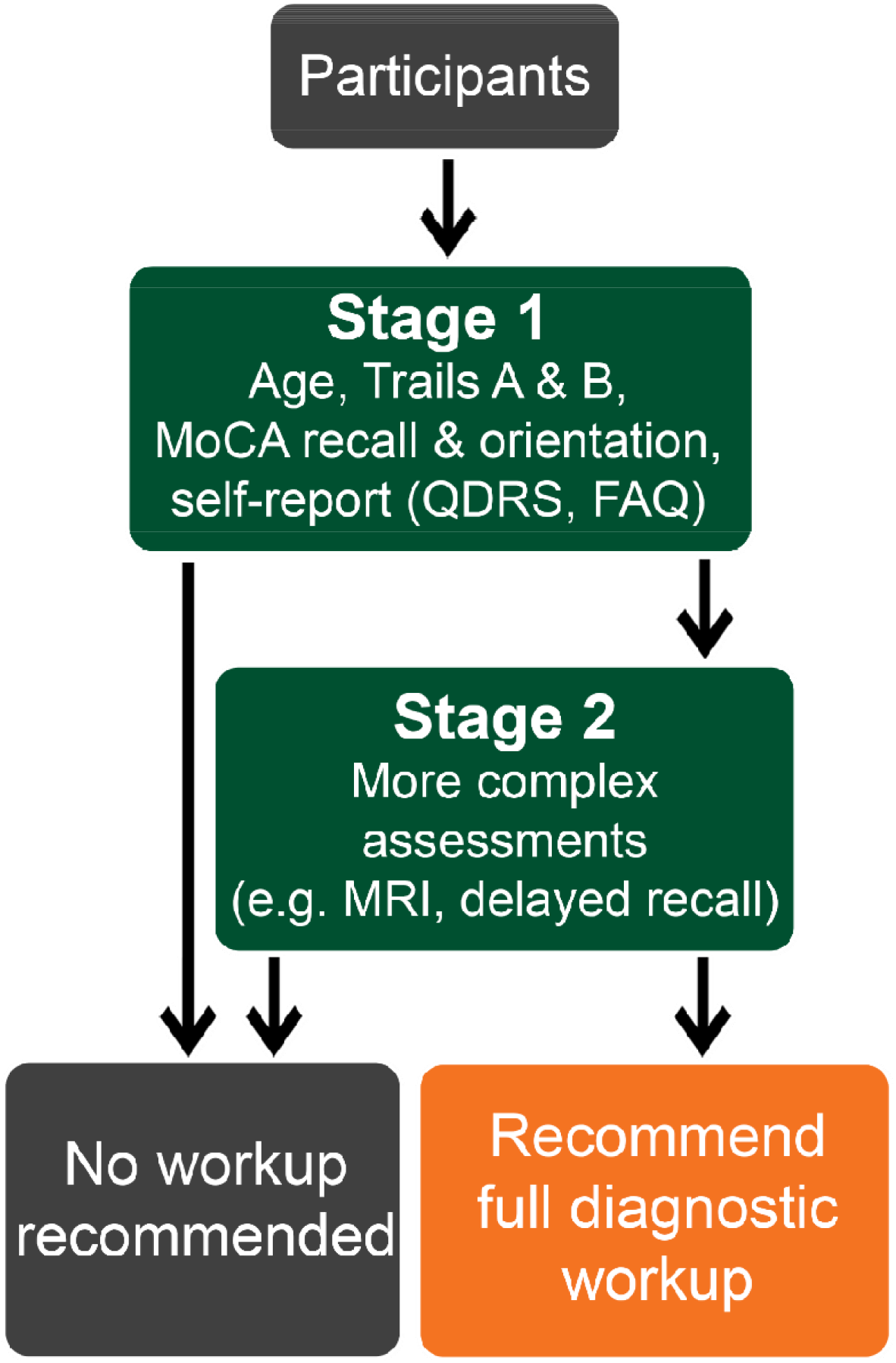
Flowchart of the hierarchical clinical decision-support system. Participants would be administered a streamlined, 5-minute assessment in Stage 1, of which the results would be entered into a clinical decision support system. The system would then recommend further testing in Stage 1 if it does not identify clear lack of impairment, followed by another assessment of patient data in the decision support system. If clear lack of impairment is again not identified, the patient would be referred for a full diagnostic workup and provided information about treatment and/or management.

### Feature selection

The *BorutaSHAP v1*.*0*.*16* [39] selection algorithm, a model-guided wrapper in Python that utilizes Shapley values to improve accuracy, was used to select the most useful and effective assessments and assessment portions (“features”) within each of the model’s two stages, using a random forest classifier (*scikit-learn v1*.*0*.*1* [40]) as its base model due to its general effectiveness. Each stage was provided a separate tailored list of features based on the intended functionality. Only one-fourth (25%) of the stratified training (ADNI) data was held for use in the feature selection process, to avoid leakage and subsequent overfitting in the model training phase.

As the primary stage is intended to identify healthy controls with minimal assessment time, only a handful of features were chosen to feed into the first feature selection algorithm. These included each component of the FAQ and its total, Trail-making Tasks A and B, the 5-word recall component of the MoCA, medical history of hypertension or stroke, educational attainment in years, and the patient’s age and sex. Self-report questionnaires that obtained the patient’s ratings of their memory, language, and attentional functioning, as well as their participation in activities within and outside the home, were also derived from ADNI’s ECog to match CCBH’s QDRS [12], including the domains 1) memory, 2) orientation, 3) judgement, 4) outside activities, 5) home activities, 6) language, and 7) attention. This questionnaire is referred to as the “QDRS-like” questionnaire.

The second stage incorporates a multi-model approach, allowing for two sets of features to each target one of the respective classes (impaired or non-impaired). Each feature selection algorithm was set to target each class, with recall (sensitivity) for that class used as the optimized parameter. In addition to the features used previous, components were considered in this stage that require extra effort or time costs. These additional features include the rest of the MoCA components and its total score, a verbal fluency task (animal naming), a verbal learning task (Rey auditory verbal learning in ADNI, Hopkins verbal learning in CCBH, adjusted for number of items) immediate and delayed recall, informant-provided versions of the QDRS-like questionnaires, and hippocampal volume as assessed by structural MRI.

The Boruta feature selection process was run a total of fifteen times: five times each for stage 1, stage 2 “impaired”, and stage 2 “non-impaired”. Features that were selected as important in at least four of the five runs per stage were selected for use in the model.

### Optimization of parameters

Each stage of the system was examined using five types of models: a logistic regression (LR), a support vector machine (SVM), a random forest (RF), a gradient-boosted machine (GBM), and a three-layer feed-forward neural network (FFNN). *Scikit-learn v1*.*0*.*1* [40] was used to create the LR, SVM, RF, and FFNN. The GBM was created using *LightGBM v3*.*3*.*2* [41], as this implementation allows for categorical variables to be accounted for and improves model time-to-fit compared to scikit-learn’s version.

*Optuna v2*.*10*.*0* [42] was used to select optimal hyperparameters for each model, leveraging its implementation of define-by-run dynamic parameter search spaces and efficient strategies for pruning. This optimization algorithm was first run to generate hyperparameters for the random forest used in the feature selection step using the same 25% of stratified training data. After feature selection was performed, *optuna* generated hyperparameters for each model based on the three feature groups: stage 1, stage 2 “non-impaired targets”, and stage 2 “impaired targets”. From the available training data, the same 30% was set aside to be used for each of the three optimization steps. Thresholding analysis was also performed to identify ideal levels for determining a classification of “impaired” or “not impaired” based on the output probabilities, also performed on the 25% stratified training data.

### Analysis

Characteristics of each dataset, as well as comparisons between datasets, were examined using either Analysis of Covariance (ANCOVA) with age as a covariate when variables were continuous, or Chi-squared tests when variables were categorical. Each model was examined for sensitivity, specificity, positive predictive value (PPV), negative predictive value (NPV), and area under the receiver-operating characteristic curve (AUC).

## RESULTS

### Participant characteristics

The sample from ADNI was comprised of 1181 participants. There were similar numbers of men (594) and women (587) in the ADNI dataset, with a significantly greater proportion of women in the HC group (57.8%) than in the MCI (44.3%) and ADRD (40.1%) groups (χ^2^(2, 1181) = 11.51, *p* < 0.001). The mean age for the ADNI sample was 72.3±7.2y. ADNI’s ADRD group (75.1±8.2) was significantly older than the HC (72.0±6.2y) and MCI (71.9±7.6y) groups (*F*(2,1178)=12.2, *p*<.001).

The sample from CCBH had 127 participants. There were more women (75) then men (52) in the CCBH dataset. While there were more women in CCBH’s HC group (75.6%) than the MCI (50.8%) and ADRD (51.8%) groups, the difference was not significant. The mean age of the CCBH sample was 72.7±10.0y. The HC group (67.6±9.1y) was significantly youngest, and the ADRD group (79.6±8.7y) was significantly oldest, with the MCI group (73.0±9.2y) significantly different from HC and ADRD (*F*(2,124)=14.4, *p*<.001).

The Clinical Dementia Rating (CDR) sum of boxes scores (CDR-SB) were not significantly different between the ADNI and CCBH samples. Non-impaired participants in ADNI had a mean CDR-SB of 0.0±0.1, and in CCBH 0.1±0.2. Impaired participants had a CDR-SB of 2.1±1.6 in ADNI and 2.2±1.8 in CCBH. Additional differences between dementia severities within each dataset can be found in **Table 1**, and comparisons between ADNI and CCBH can be found in **Table 2**.

**Table 1.** Subject Characteristics

| *Training Set (Alzheimer's Disease Neuroimaging Initiative)* |  |  |  |  |
| --- | --- | --- | --- | --- |
|  | **HC**<br>**N=517** | **MCI**<br>**N=522** | **ADRD**<br>**N=142** | **p-value** |
| Age | 72.0 (6.2) | 71.9 (7.6) | 75.1 (8.2) | **<.001** |
| Sex (% Female) | 57.8% | 44.3% | 40.1% | **<.001** |
| Years of Education | 16.7 (2.4) | 16.2 (2.6) | 15.9 (2.7) | **<.001** |
| FAQ total | 0.9 (2.5) | 6.8 (8.2) | 26.6 (11.6) | **<.001** |
| MoCA total | 25.9 (2.6) | 23.2 (3.2) | 17.6 (3.9) | **<.001** |
| Verbal fluency | 21.4 (5.5) | 18.1 (5.0) | 12.9 (5.0) | **<.001** |
| Verbal learning – Immediate | 52.9% (13.6%) | 42.5%<br>(13.1%) | 28.1% (9.0%) | **<.001** |
| Verbal learning – Delayed | 52.7% (27.2%) | 31.9%<br>(27.1%) | 3.9%<br>(8.5%) | **<.001** |
| Trailmaking Task – A | 32.1 (10.4) | 38.9 (16.8) | 57.6 (31.2) | **<.001** |
| Trailmaking Task – B | 76.3 (32.1) | 98.7 (40.4) | 146.5 (41.1) | **<.001** |
| CDR Sum of Boxes | 0.0 (0.1) | 1.4 (0.9) | 4.4 (1.6) | **<.001** |
| Hippocampal volume | 7.6 (0.9) | 7.1 (1.1) | 5.8 (1.0) | **<.001** |
| *Testing Set (Comprehensive Center for Brain Health)* |  |  |  |  |
|  | **HC**<br>**N=41** | **MCI**<br>**N=59** | **ADRD**<br>**N=27** | **p-value** |
| Age | 67.6 (9.1) | 73.0 (9.2) | 79.6 (8.7) | **<.001** |
| Sex (% Female) | 75.6% | 50.8% | 51.9% | .064 |
| Years of Education | 15.8 (2.1) | 16.0 (2.6) | 15.1 (2.5) | .475 |
| FAQ total | 0.1 (0.5) | 2.4 (3.8) | 8.7 (5.3) | **<.001** |
| MoCA total | 26.4 (2.6) | 23.2 (3.1) | 15.7 (4.0) | **<.001** |
| Verbal fluency | 20.8 (4.9) | 17.8 (4.4) | 9.7 (3.9) | **<.001** |
| Verbal learning – Immediate | 67.4% (12.1%) | 49.5%<br>(10.3%) | 27.2%<br>(10.6%) | **<.001** |
| Verbal learning – Delayed | 80.1% (15.2%) | 44.9%<br>(24.4%) | 9.0% (13.3%) | **<.001** |
| Trailmaking Task – A | 29.6 (11.8) | 34.8 (11.9) | 61.8 (28.9) | **<.001** |
| Trailmaking Task – B | 70.6 (22.7) | 93.6 (41.7) | 128.1 (48.9) | **.002** |
| CDR Sum of Boxes | 0.1 (0.2) | 1.4 (0.9) | 4.2 (1.6) | **<.001** |
| Hippocampal Volume | 6.7 (0.3) | 6.6 (0.5) | 6.4 (0.7) | .407 |
| M(SD) |  |  |  |  |
| **Bold** indicates significantly greater value |  |  |  |  |
| HC = Healthy Control; MCI = Mild Cognitive Impairment; ADRD = Alzheimer's Disease and Related Dementias; FAQ = Functional Activities Questionnaire; MoCA = Montreal Cognitive Assessment; CDR = Clinical Dementia Rating |  |  |  |  |

**Table 2.** Comparison between ADNI and CCBH

|  | Not Impaired |  | Impaired |  |
| --- | --- | --- | --- | --- |
|  | ADNI<br>N = 517 | CCBH<br>N = 41 | ADNI<br>N = 664 | CCBH<br>N = 86 |
| Age | **72.0 (6.2)** *** | 67.6 (9.1) | 72.6 (7.8) | **75.1 (9.5)** ** |
| Sex (% Female) | 57.8% | **75.6%** * | 43.4% | 51.2% |
| Years of Education | **16.7 (2.4)** * | 15.8 (2.1) | 16.1 (2.7) | 15.7 (2.6) |
| FAQ total | 0.9 (2.5) | 0.1 (0.5) | **11.0 (12.1)** *** | 4.4 (5.2) |
| MoCA total | 25.9 (2.6) | 26.4 (2.6) | 22.0 (4.1) | 20.9 (4.9) |
| Verbal fluency | 21.4 (5.5) | 20.8 (4.9) | **17.0 (5.5)** * | 15.3 (5.7) |
| Verbal learning – Immediate | 52.9%<br>(13.6%) | **67.4%** ***<br>**(12.1%)** | 39.4%<br>(13.7%) | **42.5%** **<br>**(14.7%)** |
| Verbal learning – Delayed | 52.7%<br>(27.2%) | **80.1%** ***<br>**(15.2%)** | 25.9%<br>(26.9%) | **33.6%** **<br>**(27.2%)** |
| Trailmaking Task – A | 32.1 (10.4) | 29.6 (11.8) | 42.9 (22.1) | 43.3 (22.6) |
| Trailmaking Task – B | 76.3 (32.1) | 70.6 (22.7) | 108.9 (45.0) | 104.4 (46.6) |
| CDR Sum of Boxes | 0.0 (0.1) | 0.1 (0.2) | 2.1 (1.6) | 2.2 (1.8) |
| Hippocampal Volume | **7.6 (0.9)** *** | 6.7 (0.3) | 6.8 (1.2) | 6.5 (0.6) |
| **Bold** indicates significantly greater between ADNI and CCBH<br>* = p < .05; ** = p < .01; *** = p < .001<br>ADNI = Alzheimer’s Disease Neuroimaging Initiative; CCBH = Comprehensive Center for Brain Health; FAQ = Functional Activities Questionnaire; MoCA = Montreal Cognitive Assessment; CDR = Clinical Dementia Rating |  |  |  |  |

### Primary Stage

Out of the 28 features available, the Boruta feature selection algorithm identified ten total features for the primary screening stage: the memory and language components of the QDRS-like self-report questionnaire, the “preparing paperwork” and “remembering appointments/occasions” questions of the FAQ, Trails A and B, the five-word recall and orientation components of the MoCA, the participant’s age, and the sum total of the seven QDRS-like components (**Table 3**).

**Table 3.** List of features and stage designation

| **Feature** | **Stages** | **Stage 2 Target** |
| --- | --- | --- |
| Age | Both Stages | All |
| MoCA 5-word recall | Both Stages | All |
| MoCA orientation | Both Stages | Impaired |
| MoCA Total | Stage 2 only | All |
| Trail-making A | Both Stages | Non-impaired |
| Trail-making B | Both Stages | Impaired |
| QDRS-like Memory (Self) | Both Stages | All |
| QDRS-like Language (Self) | Both Stages | All |
| QDRS-like Total (Self) | Both Stages | All |
| QDRS-like Memory (Informant) | Stage 2 only | All |
| QDRS-like Language (Informant) | Stage 2 only | All |
| QDRS-like Attention (Informant) | Stage 2 only | All |
| QDRS-like Outside (Informant) | Stage 2 only | Impaired |
| QDRS-like Total (Self + Informant) | Stage 2 only | All |
| FAQ #1 (Paying bills) | Both Stages | All |
| FAQ #2 (Paperwork) | Both Stages | All |
| FAQ #9 (Appointments) | Stage 2 only | All |
| FAQ #10 (Driving) | Stage 2 only | All |
| VLT - Delayed | Stage 2 only | All |
| Verbal Fluency (Animals) | Stage 2 only | All |
| FAQ #1 (Paying bills) | Stage 2 only | All |
| FAQ #2 (Paperwork) | Both Stages | All |
| FAQ #9 (Appointments) | Both Stages | All |
| FAQ #10 (Driving) | Stage 2 only | All |
| Hippocampal Volume | Stage 2 only | All |
| MoCA = Montreal Cognitive Assessment;<br>QDRS-like = Partially Analogous to the Quick Dementia Rating Scale;<br>FAQ = Functional Activities Questionnaire;<br>VLT = Verbal Learning Task |  |  |

After identifying ideal hyperparameters for each model type, thresholding analysis revealed that a reduced threshold of 10% (default: 50%) for positive (“impaired”) class determination was ideal for maximizing sensitivity (impaired classification) at the expense of reduced specificity (not-impaired classification). This strategy maximizes the likelihood that impaired participants are recommended for further testing.

After training each of the five models using the ADNI data, they were then tested using the held-out CCBH data. In this primary stage, the LR model had the best balance of sensitivity (96.5% or 83/86) and specificity (34.1% or 14/41), followed by the SVM (97.7% sensitivity or 84/86, and 24.4% specificity or 10/41). Area under the ROC curve analyses supported that the LR was the most balanced choice (AUC = 0.875). Using the LR as the selected model, this primary stage exhibited high diagnostic accuracy with a Positive Predictive Value (PPV) of 75.5% and a Negative Predictive Value (NPV) of 82.4%, giving a Diagnostic Odds Ratio (DOR) of 14.3 (**Table 4**).

**Table 4.** Performance of dual- and single-stage models

|  | Sensitivity | Specificity | PPV | NPV | DOR | AUC |
| --- | --- | --- | --- | --- | --- | --- |
| *Primary Stage Only Models* |  |  |  |  |  |  |
| LR | 96.5% | **34.1%** | **75.5%** | 82.4% | 14.3 | **87.5%** |
| SVM | 97.7% | 24.4% | 73.0% | 83.3% | 13.5 | 85.1% |
| GBM | 98.8% | 19.5% | 72.0% | 88.9% | **20.6** | 82.2% |
| FFNN | 98.8% | 14.6% | 70.8% | 85.7% | 14.6 | 86.1% |
| RF | **100.0%** | 0.0% | 67.7% | 0.0% | -- | 85.9% |
| *Hierarchical Two-Stage Models* |  |  |  |  |  |  |
| LR / LR | 96.5% | 36.6% | 76.1% | 83.3% | 16.0 | 87.7% |
| LR / SVM | 96.5% | 39.0% | 76.9% | 84.2% | 17.7 | 85.3% |
| LR / GBM | 95.3% | **58.6%** | **82.8%** | 85.7% | 28.9 | 89.7% |
| LR / FFNN | 91.9% | 53.7% | 80.6% | 75.9% | 13.1 | 86.5% |
| LR / RF | 91.9% | 48.8% | 79.0% | 74.1% | 10.7 | 87.9% |
| SVM / LR | 97.7% | 29.3% | 74.3% | 85.7% | 17.4 | 87.0% |
| SVM / SVM | 97.7% | 29.3% | 74.3% | 85.7% | 17.4 | 84.9% |
| SVM / GBM | 96.5% | 50.0% | 81.4% | 88.0% | 32.0 | 89.3% |
| SVM / FFNN | 91.9% | 51.2% | 79.8% | 75.0% | 11.8 | 85.9% |
| SVM / RF | 91.9% | 43.9% | 77.5% | 72.0% | 8.8 | 86.5% |
| GBM / LR | 98.8% | 41.5% | 78.0% | 94.4% | 60.2 | 88.7% |
| GBM / SVM | 98.8% | 39.0% | 77.3% | 94.1% | 54.4 | 86.0% |
| GBM / GBM | 97.7% | 46.3% | 79.2% | 90.5% | 36.3 | 89.3% |
| GBM / FFNN | 93.0% | 53.7% | 80.8% | 78.6% | 15.4 | 87.6% |
| GBM / RF | 93.0% | 41.5% | 76.9% | 73.9% | 9.4 | 87.4% |
| FFNN / LR | 98.8% | 29.3% | 74.6% | 92.3% | 35.2 | 87.8% |
| FFNN / SVM | 98.8% | 29.3% | 74.6% | 92.3% | 35.2 | 85.4% |
| FFNN / GBM | 97.7% | 53.6% | 81.6% | 91.7% | 48.6 | 89.7% |
| FFNN / FFNN | 93.0% | 51.2% | 80.0% | 77.8% | 14.0 | 86.5% |
| FFNN / RF | 93.0% | 41.5% | 76.9% | 73.9% | 9.4 | 87.1% |
| RF / LR | **100.0%** | 29.3% | 74.8% | **100.0%** | -- | 88.5% |
| RF / SVM | **100.0%** | 24.4% | 73.5% | **100.0%** | -- | 85.9% |
| RF / GBM | 98.8% | 46.3% | 79.4% | 95.0% | **73.4** | **90.1%** |
| RF / FFNN | 94.2% | 51.2% | 80.2% | 80.8% | 17.0 | 87.3% |
| RF / RF | 94.2% | 36.6% | 75.7% | 75.0% | 9.3 | 87.8% |
| *Single-Stage Models (All Features)* |  |  |  |  |  |  |
| LR | **100%** | 29.3% | 74.8% | **100%** | -- | 88.5% |
| SVM | **100%** | 24.4% | 73.5% | **100%** | -- | 85.9% |
| GBM | 98.8% | 46.3% | 79.4% | 95.0% | **73.4** | **90.1%** |
| FFNN | 94.2% | **51.2%** | **80.2%** | 80.7% | 17.0 | 87.3% |
| RF | 94.2% | 36.6% | 75.7% | 75.0% | 9.3 | 87.8% |
| **Bold** indicates highest value(s)<br>LR = Logistic Regression; SVM = Support Vector Machine; GBM = Gradient-Boosted Machine; FFNN = Feed-forward Neural Network; RF = Random Forest; PPV = Positive Predictive Value; NPV = Negative Predictive Value; AUC = Area Under the ROC Curve |  |  |  |  |  |  |

### Secondary Stage

With the inclusion of 21 additional features, bringing the total available feature count to 49, the Boruta feature selection algorithm identified two sets of features based on each of the two target classes. Both sets had 16 features in common: four questions of the FAQ (“writing checks and paying bills”, “preparing paperwork”, “remembering appointments/occasions”, and “driving or arranging transport”) plus its total score, the five-word recall component of the MoCA and its total score, the informant-provided attention, language, and memory components and the self-report memory component of the QDRS-like questionnaire plus its total, the verbal fluency task, the delayed component of the verbal learning task, the participant’s age, and hippocampal volume. Implementing the multi-model network enabled each class to be differentially targeted using features identified to best target that class, improving overall classification accuracy; when not-impaired subjects were targeted Trails A was included, and when impaired subjects were targeted Trails B, the orientation component of the MoCA, and the informant-provided “outside activities” component of the QDRS-like questionnaire were included (**Table 3**).

As in the primary stage, additional thresholding analysis was performed after identifying hyperparameters for each model on 25% of the available training data. As screening for potential impairment is the goal of this model and not diagnosis, high sensitivity at the expense of specificity was again preferred. For the RF, LR, FFNN, and SVM models, a reduced threshold of 10% for positive-class (“impaired”) determination was identified, while the GBM model performed best at a further reduced threshold of 5% for positive-class determination.

For each of the two stages, all 25 combinations of the five model types were examined (**Table 4**). Performance metrics were calculated based on all 127 subjects in the test set, with exclusions in the primary stage appended to the second stage’s outputs. As each model’s primary stage selected different subjects for use in the second stage, metrics examining the second stage only are not comparable. For example, if a subject was incorrectly classified as not-impaired in the primary stage, that erroneous classification was included when calculating sensitivity metrics for the entire two-stage model.

Sensitivity was overall high across all models, with the best performing models correctly identifying 84 out of 86 impaired participants (97.7%) (**Table 4**). Specificity was highest when a GBM was used in the secondary stage, correctly identifying 53.7%-58.6% of not-impaired participants. The LR/GBM provided the best overall combination of processing speed, sensitivity (95.3%), and specificity (58.6%), with an AUC of 0.897; the only dual-stage model that exceeded this AUC was the RF/GBM model which suffered from overfitting. The LR/GBM model also had the best Positive Predictive Value (PPV; 82.8%) and high Negative Predictive Value (NPV; 85.7%) giving a DOR of 28.9.

### Single Stage with All Features

Performance was also examined using a single-stage paradigm, where all participants were examined using only the second stage of the above model, with all available features. Across the board, performance metrics for the single-stage models were slightly better than the matched dual-stage models; for example, the GBM single-stage performed similarly to the GBM/GBM dual-stage model. The LR and SVM models achieved 100% sensitivity but with greatly reduced specificities (29% and 24%, respectively). Both the RF and FFNN models performed identically in a single stage to their paired two-stage model counterparts (RF/RF and FFNN/FFNN) (**Table 4**). However, the single stage models did not benefit from a prior screening stage and ran on all available participants.

### Misclassifications

In the well-balanced LR/GBM model, only four participants in the test set were misclassified as not impaired when they should have been screened positive. Only three were misclassified in the primary stage’s LR model, with one occurring in the secondary stage. Each of these misclassifications resembled healthy controls in all but their CDR, which ultimately guided their true diagnosis of MCI. These MCI patients had normal MoCA scores of 26.5 ± 2.6, which was similar to other HC patients scores (26.4 ± 2.6). The trail-making A scores of these misclassified patients (30.0 sec ± 3.3 sec) were more similar to HC (29.6 sec ± 11.8 sec) than MCI (34.8 sec ± 11.9 sec), and the mean score of the trail-making B task (62.5 sec ± 23.8 sec) was better than other HCs in the test set (70.6 sec ± 22.7 sec).

## DISCUSSION

This study identified two parsimonious screening stages and explored the utility of a hierarchical screening procedure to identify potential cognitive impairment and exclude patients with normal cognitive functioning, minimizing costs and reducing assessment time for these patients that may otherwise be administered additional diagnostic procedures. Optimal parameters of implementation, including the selection of machine learning model algorithms for each stage, were also explored in-depth in this study. This two-pronged approach was trained on publicly available research data (ADNI) but tested on real-world patients of a memory clinic (CCBH), exhibiting high general clinical utility especially in the 5-minute primary screening stage, and affording the ability to increase screening potential for specialists and clinical researchers in the second stage. As CDS systems become more widely integrated into clinical practice, these types of multi-tiered systems may be highly useful at ensuring those with MCI and mild ADRD are referred for formal diagnosis [20,23]. While each stage can be administered separately, enabling the quicker and simpler assessment stage to be administered in primary care and research screening contexts, a combined approach may provide a robust ability to identify very mild impairment when required.

The primary stage identified mild impairment at a high sensitivity using a delayed verbal memory component (five-word recall), a situational awareness component (orientation), a visuospatial component (trails A) and a task-switching component (trails B), along with self-report questions concerning the individual’s memory and daily functioning. Additional components in the second stage further exclude healthy participants from further testing, at the cost of increased assessment time. It should be noted that the specific assessments identified in this study may not be required for optimal utility and may be able to be modified or replaced as needed with another assessment that captures similar functioning in order to decrease costs while maintaining efficacy; for example, the task-switching component identified by Trails B may be able to be replaced with a briefer task-switching assessment, e.g., the Number-Symbol Coding Task [43]. Additionally, in the second stage, the MRI component may be able to be replaced with another measure of ADRD pathology such as fluid or PET measurements of amyloid or tau [44]. A tool that captures each of these cognitive components, but not necessarily using the exact same assessments, may thus perform similarly well as the one described in this study.

The primary stage described in this study may function effectively as an intermediary between self-report screeners, which require no clinician time except interpretation, and brief screening assessments including the full MoCA and MMSE which require at least 10 minutes. The Mini-Cog is a very brief (three minute) assessment that contains assessment components similar to that identified in the primary stage: a three-word recall delayed memory component and an executive/visuospatial clock-drawing task. However, the Mini-Cog does not incorporate task-switching, visuomotor speed, nor self-report measures.

Ultimately, while the procedure’s ability to identify current impairment was excellent, it had a tendency to misclassify cognitively normal patients as impaired even after the second stage. This was due to our prioritization of sensitivity over specificity in both stages to minimize missed identification of impairment while permitting healthy controls to be excluded following more in-depth testing; both stages are intended to screen for, not diagnose, impairment.

### Model architecture

The primary stage of the model uses quick and easily administered assessments: the trail-making test, the 5-word delayed recall and the orientation components of the MoCA, and a series of self-report questions including portions of the FAQ. This stage functions as an effective middle ground between purely questionnaire-based screening paradigms as in the QDRS [15] or FAQ [14] and brief performance assessments such as the MoCA [4] or MMSE [11]. Examining delayed memory (MoCA recall), attention (MoCA orientation), and both visuospatial and executive processing (trail-making) enables objective assessment of cognitive performance with minimal training required, and with all components able to be easily completed within five minutes; the MoCA’s five-word delayed recall component requires a five-minute delay, and both the trail-making and MoCA orientation components can be often completed within the delay portion. The self-report components of the primary stage can be completed by the participant prior to or alongside the visit, for example in the waiting room. The best performing model in this stage, the logistic regression, excluded 14 cognitively normal patients from further screening in the second stage while erroneously excluding only three borderline MCI and no AD patients; thus, sensitivity was high, but specificity was low. Performance in this stage approaches or exceeds that of similar studies that prioritize cost savings and minimize assessment time [33,36].

The model’s second stage introduces more in-depth assessments, including the verbal fluency (animal naming) task, a delayed narrative recall task, hippocampal volume from structural MRI, and informant-report questionnaires, altogether requiring a total of 30 minutes of clinician time plus the collection of the structural MRI and the involvement of an informant such as a caregiver, spouse, or family member. As the primary stage effectively identified impaired patients, this second stage functioned to better identify healthy controls for further exclusion. The best performing model in this stage, the *LightGBM* gradient-boosted machine, excluded an additional ten cognitively normal patients and only misclassified a single borderline MCI case and no ADRD patients, resulting in a total sensitivity of 95.3% and specificity of 58.6% across both stages.

Of the MCI cases that were misclassified as cognitively normal in the CCBH test set and thus excluded from further analysis, all of them exhibited normal scores on a majority of neuropsychological tests; these cases displayed normal MoCA scores, and performed better than the average non-impaired patient on the Trailmaking task versions A and B. The criteria for their diagnosis of MCI was guided by semi-structured interviews with both the patient and an informant (the Clinical Dementia Rating), revealing subtle impairment that led to a diagnosis of MCI.

### Model performance

The best performing models were found to be the combination of the LR for stage one and the GBM for stage two, as well as the model utilizing a FFNN for stage one and again using the GBM for stage two, both producing an overall AUC of 0.897. Although in the primary stage both the LR and FFNN were found to perform relatively similarly (LR AUC: 0.874, FFNN AUC: 0.861), in practice when paired with the second stage the FFNN/GBM model required 1.41 seconds to run all participants while the LR/GBM model only took 422 milliseconds to run on our machine: an improvement of over three times. The FFNN was the slowest model component overall, and despite being one of the top performers it would not be appropriate in most contexts. Further, the RF models consistently overfit in the primary stage, classifying all participants as impaired and excluding none; using a RF in the primary stage was essentially the same as not having a primary stage at all.

### Limitations

While it was a strength that the testing set was entirely separate from the training set in terms of location, participant demographics, and procedure, it was a limitation that the test set only contained a relatively small number of samples: approximately 10% of the training set. Further, the balance of impaired to non-impaired participants was not equal between the training and testing sets; the ratio in the training set was approximately 4:3 impaired to non-impaired, while the testing set was approximately 2:1 impaired to non-impaired. This was due to CCBH’s focus as a clinic open to the public, while ADNI had specific recruitment targets to fulfill their objectives of examining Alzheimer’s disease and cognitive impairment, including obtaining a representative sample of non-impaired participants. This may have also contributed to the false positives leading to overall low specificity in the test set: many controls in the test set entered CCBH with subjective complaints, potentially indicating underlying pathology that was not detected due to a lack of collection of biomarkers sensitive to prodromal impairment.

In the second stage, the use of hippocampal volume from structural MRI, as well as requiring an informant or caregiver, adds complexity and may impact the use of this stage in clinical practice and research sites. However, the removal of these components significantly impacts both sensitivity and specificity, rendering the stage less effective overall. While self-report questionnaires are fairly accurate at identifying even very mild impairment [15], informant interviews are highly useful at determining subtle impairment in daily life, an important component of MCI [16,45] and useful in ruling out non-impaired patients. Volumetric data from MRI is expensive and difficult to acquire, however future study may replace volumetric data with another biological measure, such as blood assays for dementia-related proteins (e.g., amyloid beta, phosphorylated tau) within either the primary- or secondary-stage screening procedure [46–48]. For this study, volumetric data was used both due to its availability within both datasets used as well as its validity as a neurobiological measure of ADRD.

## Conclusions

This study identified the utility of two-stage hierarchical decision support procedures and their ability to maximize screening potential while minimizing necessary costs, compared to a single model using the features of both stages. The development of the procedure revealed that a brief 5-minute assessment of delayed verbal memory, visuospatial and executive functioning, and attention along with self-report memory and IADL questions, is highly effective at identifying MCI and ADRD. Additional examination using the optional second-stage of the procedure is able to further exclude non-impaired individuals. Additional optimization and validation using more diverse populations is needed, as is exploration of a more parsimonious second stage.

## Data Availability

All data produced in the present study are available either upon reasonable request to the authors for the memory clinic data, or available online for the ADNI data

http://adni.loni.usc.edu

## ACKNOWLEDGEMENTS

Work on this study was supported by grants from the National Institute on Aging (R01 AG071514, R01 AG069765, and R01 NS101483), the Alzheimer’s Association, the McKnight Brain Research Foundation, the Harry T. Mangurian Foundation, and the Leo and Anne Albert Charitable Trust.

Data collection and sharing for this project was funded by the Alzheimer’s Disease Neuroimaging Initiative (ADNI) (National Institutes of Health Grant U01 AG024904) and DOD ADNI (Department of Defense award number W81XWH-12-2-0012). ADNI is funded by the National Institute on Aging, the National Institute of Biomedical Imaging and Bioengineering, and through generous contributions from the following: AbbVie, Alzheimer’s Association; Alzheimer’s Drug Discovery Foundation; Araclon Biotech; BioClinica, Inc.; Biogen; Bristol-Myers Squibb Company; CereSpir, Inc.; Cogstate; Eisai Inc.; Elan Pharmaceuticals, Inc.; Eli Lilly and Company; EuroImmun; F. Hoffmann-La Roche Ltd and its affiliated company Genentech, Inc.; Fujirebio; GE Healthcare; IXICO Ltd.; Janssen Alzheimer Immunotherapy Research & Development, LLC.; Johnson & Johnson Pharmaceutical Research & Development LLC.; Lumosity; Lundbeck; Merck & Co., Inc.; Meso Scale Diagnostics, LLC.; NeuroRx Research; Neurotrack Technologies; Novartis Pharmaceuticals Corporation; Pfizer Inc.; Piramal Imaging; Servier; Takeda Pharmaceutical Company; and Transition Therapeutics. The Canadian Institutes of Health Research is providing funds to support ADNI clinical sites in Canada. Private sector contributions are facilitated by the Foundation for the National Institutes of Health (www.fnih.org). The grantee organization is the Northern California Institute for Research and Education, and the study is coordinated by the Alzheimer’s Therapeutic Research Institute at the University of Southern California. ADNI data are disseminated by the Laboratory for Neuro Imaging at the University of Southern California.

## DISCLOSURE STATEMENT

The authors have no conflict of interest to report.

## Notes

### Competing Interest Statement

The authors have declared no competing interest.

### Author Declarations

Approval was granted by the University of Miami Institutional Review Board

